# Synchronized travel restrictions across cities can be effective in COVID-19 control

**DOI:** 10.1101/2020.04.02.20050781

**Authors:** Haiyan Liu, Xuemei Bai, Huanfeng Shen, Xiaoping Pang, Zeyu Liang, Yue Liu

## Abstract

The COVID-19 outbreak is under control in China. Mobility interventions, including both the Wuhan lockdown and travel restrictions in other cities, have been undertaken in China to mitigate the epidemic. However, the impact of mobility restrictions in cites outside Wuhan has not been systematically analyzed. Here we ascertain the relationships between all mobility patterns and the epidemic trajectory in Chinese cities outside Hubei Province, and we estimate the impact of local travel restrictions. We estimate local inter-city travel bans averted 22.4% (95% PI: 16.8–27.9%) more infections in the two weeks after the Wuhan lockdown, while local intra-city travel prevented 32.5% (95% PI: 18.9–46.1%) more infections in the third and fourth weeks. More synchronized implementation of mobility interventions would further decrease the number of confirmed cases in the first two weeks by 15.7% (95% PI:15.4–16.0%). This study shows synchronized travel restrictions across cities can be effective in COVID-19 control.

## Introduction

On 12 March 2020, the World Health Organization (WHO) formally declared the novel coronavirus disease 2019 (COVID-19) outbreak as a global pandemic, urging countries to take precautionary public health measures to curb its spread^1^. Until that date, the COVID-19 virus had spread to over 117 countries and areas in the world, with 125,260 confirmed cases and 4,613 reported deaths^2^. Since December 2019, China has reported a total of 80,981 infected individuals and 3,173 deaths^2^. After 11 March 2020, without considering the imported cases from other countries, there are no new confirmed cases in China except in Wuhan^3,4^; the outbreak of COVID-19 is under control in China^5^. China’s transmission control measures to stem the spread of COVID-19 may provide valuable experience for other countries.

Previous studies prove it is population mobility that accelerates the spatial spread of the epidemic, while travel restrictions could contribute to epidemic control^5–13^. On 10 January, the 2020 spring migration to celebrate Chinese New Year began – the world’s most massive annual human migration^13^. Although Wuhan, the capital city of Hubei Province, suspended all inflow and outflow public transport from 10:00 a.m. on 23 January 2020, about 5 million people had already left Wuhan before the quarantine^14^. Studies show that while the Wuhan lockdown greatly slowed the spread of COVID-19^6–10^, the population emigration figure from Wuhan highly correlated to the imported cases in other cities in China^8,11–13,15–18^.

While the impact of the population outflow from Wuhan is well established, the impact of other mobility patterns on the epidemic trajectory has not been well understood. Local population mobility for a city includes both intra-city and inter-city patterns. The inter-city mobility can be categorized into three sources, i.e., from Wuhan, from Hubei province (excluding Wuhan), and from cities outside Hubei. As nearly two-thirds of population outflow from Wuhan flooded into other cities within Hubei Province^13^, it is important to consider the population outflow from Hubei (excluding Wuhan) as a potentially significant source of epidemic transmission risk after 23 January^17^. The intra-city population movement is also an essential factor – research shows cities that introduced pre-emptive intra-city movement restrictions have 33.3% fewer confirmed cases in the first week of the epidemic outbreak compared to those that started restrictions after the emerging of confirmed cases^6^, pointing to the importance of the timing of introducing these measures. For inter-city population movement among cities outside Hubei, which are restricted after the Wuhan lockdown, different studies hold inconsistent views. On the one hand, the implementation of inter-city travel restrictions cannot significantly reduce the number of confirmed cases during the first week of city outbreaks^6^; on the other hand, the transmission model of COVID-19 cannot be accurately established without the inter-city connections^19^.

Most of these existing studies are based on one or two mobility patterns, and the overall impact of these four mobility patterns on the spread of the coronavirus is not well understood. The uncertainty of the number of confirmed cases in the early stage of the epidemic spread^5,19^, ranging from 427 (officially confirmed) to potentially over 10,000 underscores the importance of considering all four mobility patterns in the COVID-19 spread, perhaps more so than the number of confirmed cases.

In this study, we aim to investigate the relationship between population mobility, both inter-city and intra-city, and the spread of COVID-19. Based on the mobility change caused by the implementation of restrictions, we estimated the impact of local travel restrictions in cities outside Hubei (Fig. 1) – outside of the Wuhan lockdown – on control of the epidemic. Given that the three mobility patterns, except the population outflow from Wuhan, are heavily influenced by policy measures introduced by the central and local governments, such investigation is important in evaluating the effectiveness of and understanding the influence of the timing of different measures, which can in turn inform policy interventions in the future.

**Fig. 1.**
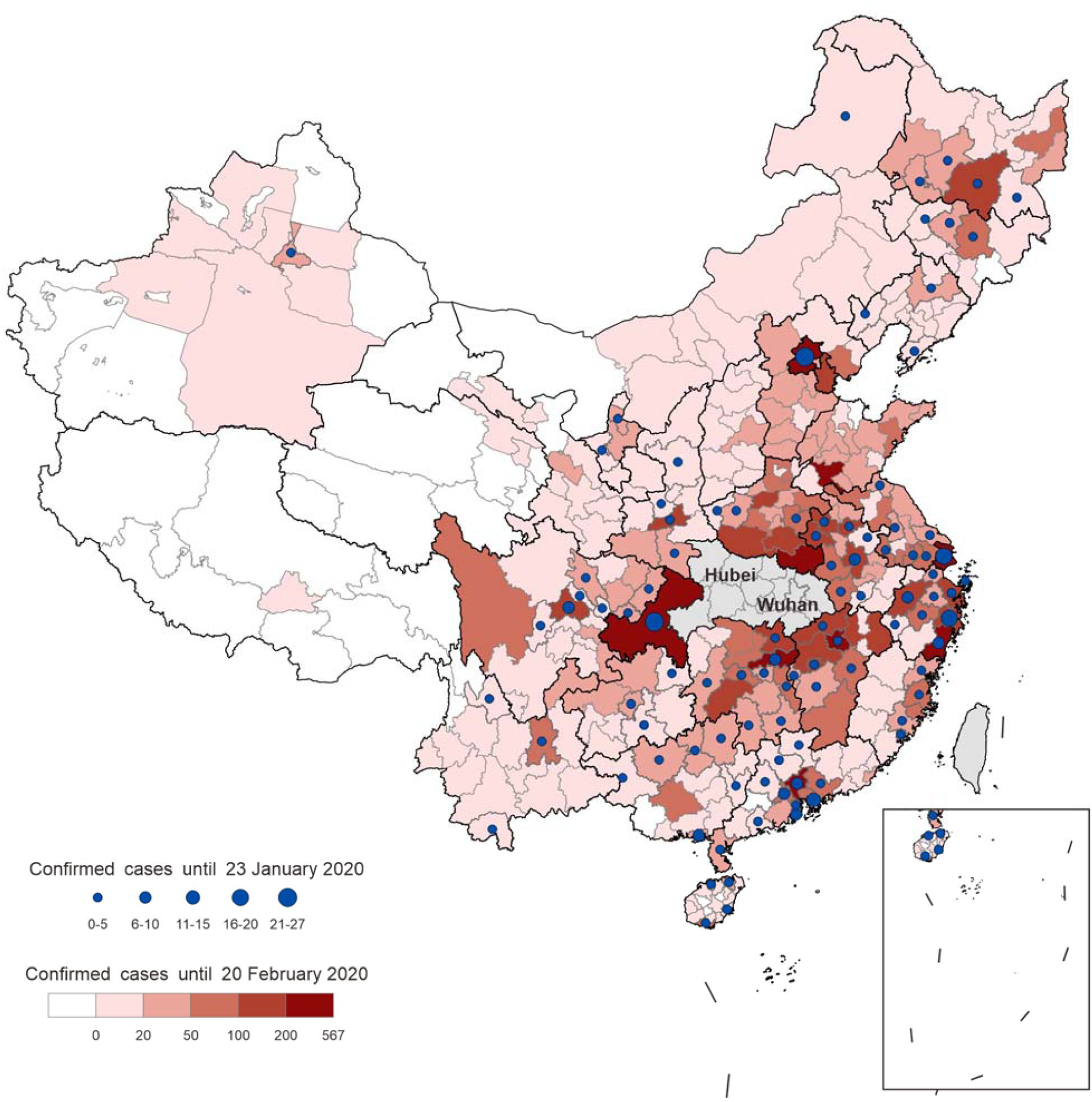
Cities outside Hubei Province and their number of cumulative confirmed cases. Here the blue circles represent the number of cumulative confirmed cases in cities outside Hubei by 23 January 2020, while cities without circles are those without initial confirmed cases until the Wuhan lockdown. The number of cumulative confirmed cases are shown in red, while the white represents the cities with no reported cases before 20 February 2020. The confirmed cases of Fushun were recorded into Shenyang, so these two cities were merged for later analysis.

## Results

### Relationship between different mobility patterns and epidemic spread

All mobility patterns are positively correlated with the COVID-19 outbreak in both Stage One and Stage Two (p<0.01) (Table 1-1 and 1-2, Supplementary Figure 1). This suggests that the following would all cause more confirmed cases in the first two weeks after the Wuhan lockdown: 1) cities with more population inflow from Wuhan and other cities in Hubei, 2) increased population migration from cities outside Hubei, and 3) more intra-city population movement. Moreover, in Stage Two when local travel bans have been fully implemented, cities with a larger number of inter-city and intra-city population movements would have more local infections.

**Table 1-1.** Pearson correlation coefficient (r) between the number of confirmed cases and population mobility data in Stage One. (Symbols used here are mentioned in Data and methods, and *i*=305.)

| r and p-value | $C_i$ | $E(w)_i$ | $E(h)_i$ | $O_i$ | $I_i$ |
| --- | --- | --- | --- | --- | --- |
| $C_i$ | 1 | 0.717* | 0.739* | 0.650* | 0.597* |
| $E(w)_i$ | 0.717* | 1 | 0.667* | 0.584* | 0.598* |
| $E(h)_i$ | 0.739* | 0.667* | 1 | 0.595* | 0.506* |
| $O_i$ | 0.650* | 0.584* | 0.595* | 1 | 0.812* |
| $I_i$ | 0.597* | 0.598* | 0.506* | 0.812* | 1 |

\* indicates significance at the 1% level

**Table 1-2.** Pearson correlation coefficient (r) between the number of confirmed cases and population mobility data in Stage Two. (Symbols used here are mentioned in Data and methods, and *i*=250.)

| r and p-value | $C'_i$ | $O_i$ | $I_i$ |
| --- | --- | --- | --- |
| $C'_i$ | 1 | 0.456* | 0.442* |
| $O_i$ | 0.456* | 1 | 0.465* |
| $I_i$ | 0.442* | 0.465* | 1 |

\* indicates significance at the 1% level

The correlation coefficients between the same mobility pattern and the number of confirmed cases are different in Stages One and Two. In Stage One, the coefficients between the population inflow from Wuhan and other cities in Hubei and the number of confirmed cases (r=0.717 and 0.739 respectively, p<0.01) are higher than those of population migration from cities outside Hubei and than intra-city population movement (r=0.650 and 0.597 respectively, p<0.01), indicating the leading role of imported cases in this time period. In Stage Two, the correlations between mobility patterns and the number of confirmed cases (r<0.5, p<0.01) weaken compared to those in Stage One (r>0.55, p<0.01), suggesting the effects of mobility on epidemic spread might be reduced due to local travel restrictions. Meanwhile, the relationship between population migration from cities outside Hubei and intra-city population movement changes from highly correlated (r=0.812, p<0.01) to moderately correlated (r=0.465, p<0.01), implying the different strictness of inter-city and intra-city travel bans.

The best-fitting linear regression models for epidemic development in both Stage One and Stage Two are listed in Table 2. In Stage One, three inter-city mobility patterns, together with the number of initial confirmed cases, could explain 73.3% of the inter-city differences in newly reported infections in cities outside Hubei. The intra-city population movement figure is highly correlated with the inflow from cities outside Hubei (r=0.812, p<0.01), but its impact on the local epidemic development is less than the inter-city scenario. This result implies the existence of imported cases from cities outside Hubei. In Stage Two, intra-city population movement and the number of initial confirmed cases could explain 51.0% of the inter-city differences in newly reported infections in cities outside Hubei. The impact of inter-city population movement is not significant in this period.

**Table 2.** Impact of the different mobility patterns evaluated by linear regression models.

|  | Stage One |  |  |  |  |  |  |  |  | Stage Two |  |  |
| --- | --- | --- | --- | --- | --- | --- | --- | --- | --- | --- | --- | --- |
| | $C_i > 0$ ($i=305$) | | | $C_{0i}=0$ ($i=206$) | | | $C_{0i}>0$ ($i=99$) | | | $C'_i > 0$ ($i=250$) | | |
| Independent variables | βs | 95% CI | p-value | βs | 95% CI | p-value | βs | 95% CI | p-value | $\beta$'s | 95% CI | p-value |
| $\varepsilon$ | 4.07 | (-1.05, 9.20) | 0.12 | 1.51 | (-0.97, 4.00) | 0.23 | 11.17 | (0.32, 22.03) | 0.04 | 1.03 | (-2.67, 4.74) | 0.58 |
| $E(w)_i$ | 35.39 | (26.72, 44.07) | <0.01 | 35.51 | (31.50, 39.51) | <0.01 | 39.48 | (14.08, 64.88) | <0.01 | / | / | / |
| $E(h)_i$ | 12.71 | (6.79, 18.62) | <0.01 | 4.75 | (1.34, 8.15) | <0.01 | 19.66 | (6.14, 33.17) | <0.01 | / | / | / |
| $O_i$ | 0.10 | (0.03, 0.16) | <0.01 | 0.07 | (0.01, 0.13) | <0.01 | / | / | / | / | / | / |
| $I_i$ | / | / | / | 0.002 | (0.001, 0.004) | <0.01 | / | / | / | 0.003 | (0.001, 0.006) | <0.01 |
| $C_{0i}$ | 6.35 | (5.01, 7.69) | <0.01 | / | / | / | 5.96 | (3.24, 8.68) | <0.01 | 0.26 | (0.22, 0.31) | <0.01 |
| Adjusted R<sup>2</sup> | 0.733 |  |  | 0.808 |  |  | 0.680 |  |  | 0.510 |  |  |

In Stage One, the epidemic spread in cities outside Hubei, with or without initial confirmed cases, is significantly influenced by population inflow from Hubei, including from Wuhan and from other cities in Hubei. Meanwhile, the inter-city population movement from cities outside Hubei and intra-city population movement have varied impact. For cities without initial confirmed cases, both inter-city and intra-city population movements significantly influenced the epidemic development. In other words, four kinds of mobility patterns jointly affected the epidemic spread in these cities. For cities with initial confirmed cases, both inter-city (from cities outside Hubei) and intra-city population movement have no significant impact on their epidemic development. Two reasons might cause this. First, the directly imported cases from Hubei took up the majority of reported cases in this stage, namely the significance of population inflow from Hubei subjugating others. The cities with initial confirmed cases are the leading destinations for population outflow from Hubei. These cities, making up 32.6% of cities outside Hubei, accommodated 57.9% of population outflow from Wuhan before its closure and 68.1% of population outflow from Hubei (excluding Wuhan) after the Wuhan lockdown. Second, in these cities both population inflow from cities outside Hubei and intra-city population movement are highly correlated with the population inflow from Wuhan (r=0.700 and 0.748 respectively, p<0.01). The inter-city and intra-city mobility data might be excluded from the best-fit model due to multicollinearity.

### The impact of local travel restrictions in cities outside Hubei on the epidemic spread

Local travel restrictions in cities outside Hubei have contributed to the epidemic control (Table 3). Using the best-fitting model, we estimated that if there were no travel restrictions in cites outside Hubei in the first week after the Wuhan lockdown (Fig. 2b), the number of infections in Stage One would have increased by 1,960 (95% PI: 1,474–2,447), which equates to 22.4% (95% PI: 16.8–27.9%) of the confirmed cases. Most of the growth would have happened in the cities with initial confirmed cases. For these cities, the growth in numbers would be 1,403 (95% PI: 851–1,954), which is 26.1% (95% PI: 15.8–36.3%) of confirmed cases in these cities. An increase of 579 (95% PI: 441–717) cases is expected to have appeared in cities without initial confirmed cases up until 23 January, which translates to 17.1% (95% PI: 13.0–21.2%) of their confirmed cases.

**Table 3.**
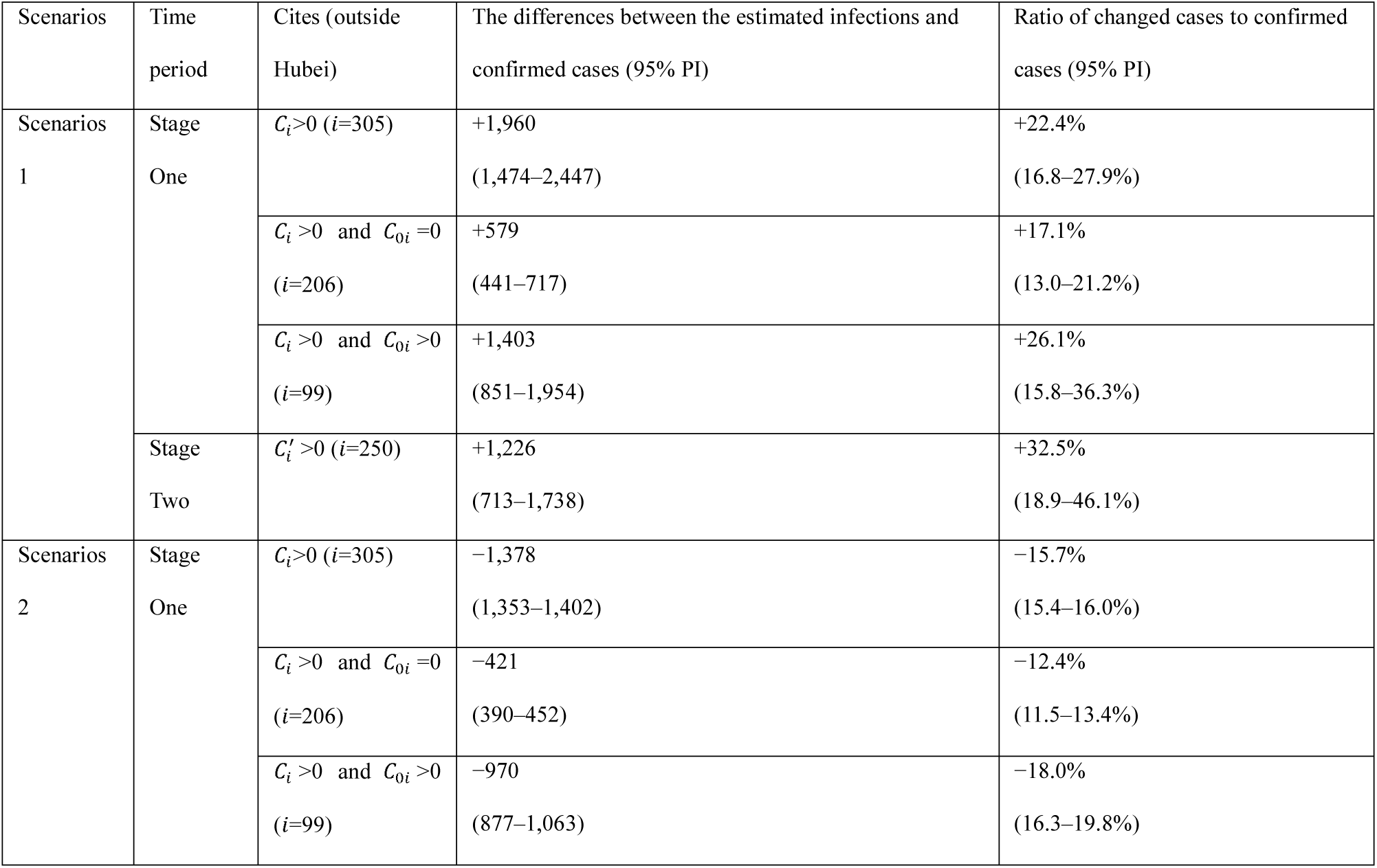
The differences between the estimated number of infections and actual confirmed cases in cities outside Hubei under different scenarios.

**Fig. 2.**
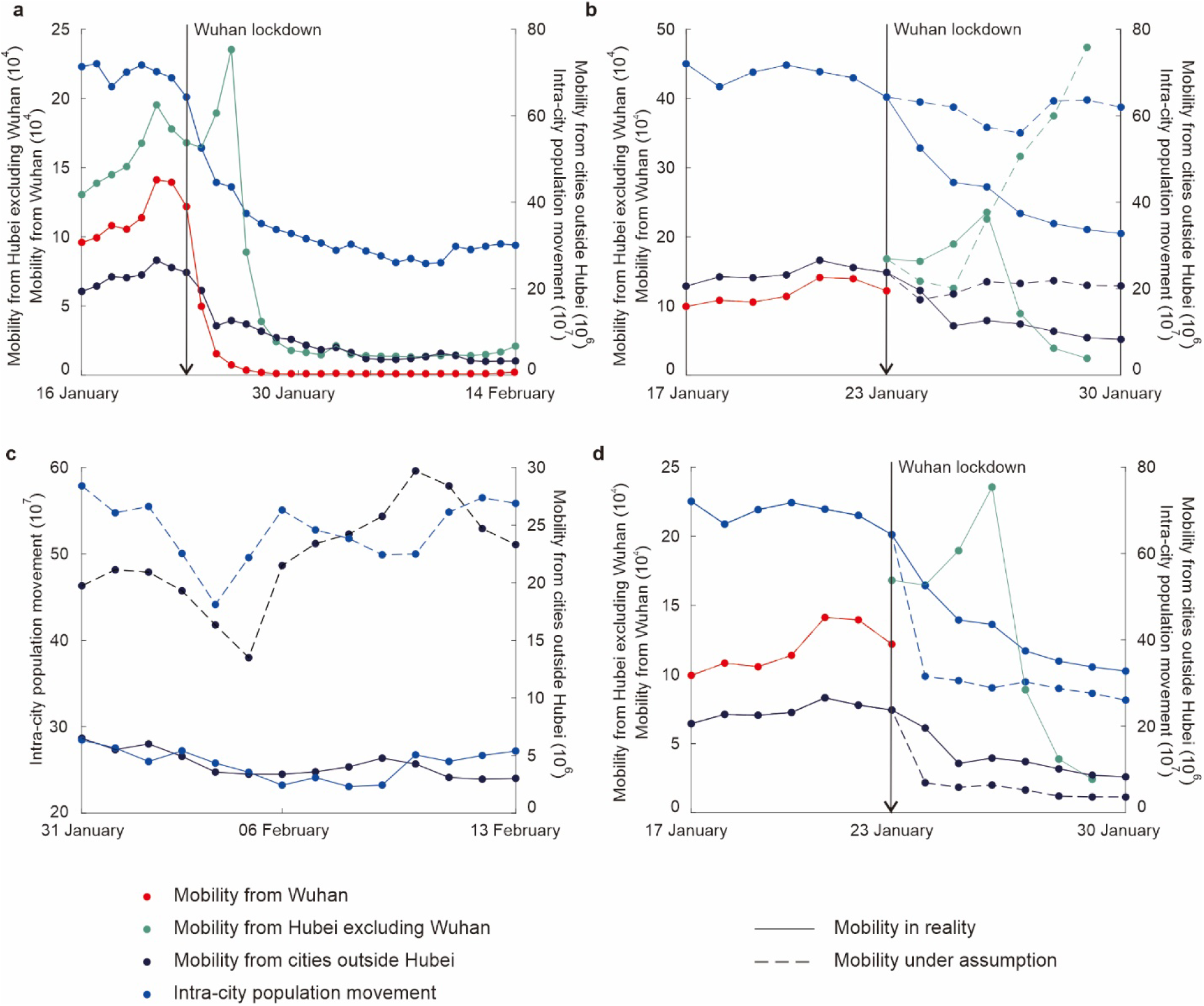
Mobility change under different scenarios. **a**. Daily sum of mobility data in cities with confirmed cases in Stage One or Stage Two from 16 January–14 February 2020. **b**. Data of four mobility patterns that correlate with the cumulative number of confirmed cases in Stage One and their estimated value without local travel restrictions excluding the Wuhan lockdown. Each point is the daily sum of one mobility pattern in the 305 cities having confirmed cases in Stage One. **c**. Data of two mobility patterns that correlate with the cumulative number of confirmed cases in Stage Two and their estimated value without local travel restrictions. Each point is the daily sum of one mobility pattern in the 250 cities having confirmed cases in Stage Two. **d**. Data of four mobility patterns that correlate with the cumulative number of confirmed cases in Stage One and their estimated value with synchronized travel restrictions across cities. For the estimated value of population outflow from Hubei excluding Wuhan, we used one-day data on 23 January 2020. Each point is the daily sum of one mobility pattern in the 305 cities having confirmed cases in Stage One.

Most of the estimated increase seen during Stage One is due to the population inflow from Hubei after the Wuhan lockdown. If we saw only the population outflow from Hubei prohibited after the Wuhan lockdown, i.e., the inter-city and intra-city population movement were not restricted in cities outside Hubei, the estimated increase in the number of confirmed cases would be 1,384 (95% PI: 1,016–1,752) (Supplementary Table 2). Thus, more than two-thirds of the estimated increase in infections is due to the population outflow from Hubei (excluding Wuhan) if there were no travel bans except the Wuhan lockdown.

An intra-city travel ban plays a significant role in preventing local infections in Stage Two (Table 3). There were only four cities without infections in Stage One but they all reported infections in Stage Two. If there is no intra-city travel restriction (Fig. 2c), the confirmed infections are estimated to increase by 1,226 (95% PI: 713–1,738), which is 32.5% (95% PI: 18.9–46.1%) of confirmed cases in these cities.

Although the travel restrictions in Chinese cities outside Hubei have played essential roles in the epidemic control, our results suggest that more timely travel restrictions after the Wuhan lockdown could better control the spread of the virus. If all cities outside Hubei imposed their inter-city travel bans, including to and from Hubei, and intra-city travel bans at the same time as the Wuhan lockdown (Fig. 2d), these cities would report 1,378 (95% PI: 1,353–1,402) fewer cases than those confirmed in Stage One, which is 15.7% (95% PI: 15.4–16.0%) (Table 3). Furthermore, if only Hubei was shut down at the same time as Wuhan, the confirmed cases would reduce by 918 (95% PI: 896–940) in the following two weeks, which represents 10.5% (95% PI: 10.2–10.7%) of confirmed cases (Supplementary Table 2).

## Discussion

Mobility control measures are of crucial importance for public health planning during the outbreak of COVID-19^7^. In this study, we explore the relationships between mobility and epidemic spread and estimate the impact of local travel restrictions on epidemic control. Our findings suggest that the travel bans imposed by cities outside Hubei have prevented 1,960 (95% PI: 1,474–2,447) confirmed cases, representing 22.4% (95% PI: 16.8–27.9%) of observed cases, in the two weeks after the Wuhan lockdown. More timely travel bans would further decrease the number of confirmed cases in the same period by 15.7% (95% PI:15.4%–16.0%) or 1,378 (95% PI: 1,353–1,402) cases. Therefore, besides the lockdown of one epidemic city, the timely implementation of travel restrictions – including both inter-city and intra-city travel bans – in other cities can effectively control the COVID-19 outbreak.

If the whole of Hubei province was not quarantined after the Wuhan lockdown, further national seeding and subsequent infections might have become inevitable. By 12 March 2020, Hubei (excluding Wuhan) has more confirmed cases (17,795) than China excluding Hubei (13,032)^24^. Our results suggest that in cities outside Hubei, the travel restriction of Hubei (excluding Wuhan) is more effective than other inter-city and intra-city travel bans in controlling the development of the epidemic in the two weeks after the Wuhan lockdown. On January 26, all airports and railway stations in Hubei were closed^25^. Before January 30, all other provinces in mainland China suspended their inter-provincial road transport to and from Hubei (Supplementary Table 1), quarantining the whole of Hubei Province. Although these travel restrictions reduced the number of confirmed cases in cities outside Hubei by 15.8% (95% PI: 11.6%–20.0%) of reported cases in the two weeks after the Wuhan lockdown, another 10.5% (95% PI: 10.2–10.7%) might have been prevented by a more timely quarantine of the whole Hubei province (Supplementary Table 2). This highlights the importance of timely and coordinated response across localities in epidemic mitigation.

For cities with and without initial confirmed cases by the time of the Wuhan lockdown, their local epidemics might be dominated by different population mobility patterns. Most cities reporting confirmed cases before the Wuhan shutdown are the leading destinations for population outflow from Wuhan and other cities in Hubei. Cutting off the inter-provincial traffic from Hubei could protect them to a great extent. For cities without reported infections before the Wuhan lockdown, the preventive prohibition of both inter-city and intra-city population movement is essential to their epidemic control. The prohibition of inter-city population movement from cities outside Hubei and intra-city population movement prevented 405 (95% PI: 342–468) more infections in Stage One (Supplementary Table 2), occupying on average 69.9% of the number of preventions by all local travel bans. The travel controls in cities without initial confirmed cases tend to be relatively late or looser than those in cities with initial infections. From 24–30 January 2020, the population inflow from cities outside Hubei and intra-city population movement in cities without initial confirmed cases decreased on average to 64.4% and 72.6% of those in the same period of 2019, while the percentages are on average 59.2% and 65.2% for cities with initial confirmed cases (Fig. 2b). The local travel restrictions were necessary and practical for cities without initial confirmed cases, even if they were not that strict.

Different mobility patterns influenced the COVID-19 spread in different periods. Our results show in the early stage of epidemic development it is the inter-city mobility – including from Wuhan, from Hubei excluding Wuhan, and from other cities outside Hubei – that promotes the spatial spread of the coronavirus. During the implementation of local travel bans, the inter-city population movement was more strictly restricted than the intra-city traveling, which limited the further spatial spread of coronavirus in Stage Two. The intra-city travel bans cause the intra-city population movement from 31 January–13 February (after the public holiday of the spring festival) to decrease to 50.1% of that in the previous year (Fig. 2c), ranging from 23.7–87.8% across cities. For these cities, the inter-city population movement on average decreased to 15.8% of that in the previous year (Fig. 2c), ranging from 2.8–55.7%. After the quarantine of the whole Hubei and the prohibition of inter-city transport, the importance of restricting the intra-city population movement is highlighted. The confirmed cases could increase by 32.5% (95% PI: 18.9–46.1%) of reported cases in cities outside Hubei if there were no local intra-city travel bans, suggesting the intra-city travel restrictions played vital roles in the epidemic control in China.

It is worth noting that China has implemented many non-pharmacological interventions, not limited to these travel restrictions. Source control measures, like isolating people with the virus, monitoring or quarantining symptoms of healthy contacts, requiring masks for individuals in all public places, etc., have been introduced to reduce potential secondary infections^26,27^. It is the intensive source control that reduces new local infections. The contribution of population movement from Wuhan or Hubei to subsequent epidemic development might also be dampened due to the implementation of source control measures^7^. Without the implementation of combined source control measures, our study, at least in the model of Stage Two, might have different results.

Our study quantified the relationships between mobility patterns and epidemic trajectory in China and highlighted the importance of synchronized travel restrictions across cities. Several policy implications can be drawn. First, the geographical extension of the quarantine should be carefully considered by the government before the official announcement. In the early stage of epidemic development, there might be a non-negligible number of infected but undetected people in the areas that have geographical connections and frequent traffic with the epidemic-stricken ones, like other cities in Hubei. In particular, there are many asymptomatic virus carriers who are highly contagious^23,28^. On the other hand, the expansion of the quarantined area would also bring substantial economic losses. Whether to include farther hinterlands, and if so to what extent, will need to be considered carefully in making the quarantine decision. Second, our results show the importance of timely and active local countermeasures by cities outside of the epicenter. While actual travel control measures may differ across cities, simply compressing the time gap between Wuhan and other cities could further reduce the COVID-19 outbreak. It is important for countries and governments to impose timely interventions to combat the pandemic.

## Data and methods

### Number of confirmed cases

We collected the number of laboratory-confirmed cases from daily official reports from the health commissions of the 34 provincial-level administrative units from 23 January 2020. The provincial reports include the total instances as well as the breakdown for cities. The city-level data we used includes 367 cities, i.e., 4 cities directly under the central government, 333 prefecture-level cities, and 29 county-level cities directly under the provincial government, covering all the areas of mainland China. A total of 350 cities are from outside Hubei.

### Population mobility data

To capture population movement among and within cities, we derived the mobility data from Baidu Qianxi^20^. The data is calculated and analyzed from the location-based service (LBS) of both Baidu Map and a flight-path monitoring app, Baidu Tianyan. Data from both 2019 and 2020 were acquired, aligned with the Chinese lunar calendar, from the start of spring migration (21 January 2019 and 10 January 2020).

The data of population inflow from Wuhan, from Hubei excluding Wuhan, and from cities outside Hubei were all calculated based on the Baidu Mobility Index. The Baidu Mobility Index records daily outflow and inflow to and from each of the 367 cities, which is comparable among cities. We assumed there were 5 million people outflowed from Wuhan between the start of the spring emigration and the Wuhan lockdown^14^, and scaled the Index to approximate values of population size. For each day, the top 100 destination cities for population outflow from Wuhan and other cities in Hubei were recorded. We believe the data are representative of population outflow from Wuhan and from Hubei excluding Wuhan^19^. For the population inflow from cities outside Hubei, we used the overall daily inflow values of each city and subtracted the population migration figure from all cities in Hubei.

The data of the intra-city population movement were counted based on the Baidu Intra-city Mobility Index. The Intra-city Mobility Index, ranging from 0.3–8.0, reflects the proportion of people traveling within cities in the resident population^20^. We used the 0.1 times of the daily Intra-city Mobility Index, ranging from 3.0–80.0%, multiplied with the value of permanent population as the proxy daily population intra-city travel data. The data of the permanent population at the end of 2018 was retrieved from the statistical yearbook of provinces and cities.

### Identifying the relationships between different mobility patterns and epidemic spread

The imported cases and local infected ones might be caused by different mobility patterns. Thus, we used the cumulative confirmed cases of two time periods in cities outside Hubei to study their relationship with different mobility data.

The cumulative confirmed cases in two periods were computed to simulate the epidemic spread at different stages. The first period, referred to hereafter as Stage One, was the first two weeks after the Wuhan lockdown, from 24 January–6 February 2020. Since the incubation period for COVID-19 usually ranged from 1–14 days, the majority of imported cases would be identified in this period. The second period was the two weeks after Stage One, referred to hereafter as Stage Two, from 7–20 February 2020. Most of the confirmed cases in this period should be infected in cities outside Hubei.

The ratio of new confirmed cases in the population outflow was typically used to estimate the virus transmission risk of population outflow^17^. However, the epidemic outbreak might be underestimated at the early stage due to the lack of attention or detection ability. Until the Wuhan shutdown, only 427 cases were confirmed as positive for COVID-19, while the 86% of all infections might be undocumented.^19^ Facing such a big gap, we investigate the relationship between mobility and epidemic spread without considering the proportion of the number of confirmed cases in population migration.

To identify the impact of different mobility patterns on the epidemic spread, we used linear regression models^6^ to assess the relationship between population mobility data and the confirmed cases. In Stage One, the confirmed cases could be imported from Hubei (both Wuhan and cities outside Wuhan), from cities outside Hubei, or from the locality. Therefore, the analysis was performed using the model: 

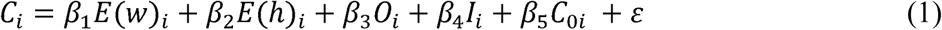

Where *C*_*i*_ is the number of cumulative confirmed cases in Stage One of city *i*; *E*(*w*)_*i*_ is the number of population inflow from Wuhan before its lockdown; *E*(*h*)_*i*_ is the number of population inflow from Hubei excluding Wuhan;*O*_*i*_ represents inter-city population movement, using the number of population inflow from cities outside Hubei; *I*_*i*_ is the number of intra-city population movement; *C*_0*i*_ is the number of initial confirmed cases, which is the cumulative confirmed cases until January 23 of city *i*; *β*_1_, *β*_2_, *β*_3_, *β*_4_, and are the regression coefficients; *ε* and is the constant coefficient that reflects information residue.

Since the mean incubation period of COVID-19 was 5.2 days (95% confidence interval [CI], 4.1–7.0)^21^, changing across studies^7,22,23^, we used the mobility data for one week before the reported day in the model. The data of *E*(*w*)_*i*_ was from 17–23 January, while the data of and were all from 17–30 January. As for *E*(*h*)_*i*_, we used the data after the Wuhan lockdown because it was not the main risk of transmission before that^17^. Thus, the data of *E*(*h*)_*i*_ were from 23 January to the suspension of all inter-provincial transport to and from Hubei, 29 January (Supplementary Table 1).

As for Stage Two, the imported cases from Hubei were not considered: 

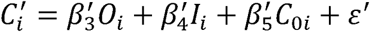

Where 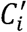 is the number of cumulative confirmed cases in Stage Two of city *i*; *O*_*i*_ is the number of inter-city population movement from 31 January–13 February; *I*_*i*_ is the number of intra-city population movement 31 January–13 February; and *C*_0*i*_ is the number of initial confirmed cases until 6 February.

The statistical analysis included two steps. First, Pearson correlation analysis was applied to check whether different mobility patterns were correlated with the spread of COVID-19 in cities outside Hubei in two time periods. Correlated data would be introduced to the following linear regression. Second, stepwise multivariate linear regressions were built to explore the explanatory capacity of mobility data to the number of confirmed cases. A model with the highest adjusted R^2^ was taken as the best-fit scenario. Cities without confirmed cases until the end of each stage were excluded from the study. Besides, for all the cities with confirmed cases after the Wuhan lockdown (*C*_*i*_ > 0 or 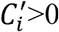), we also generated models for cities with (*C*_0*i*_=0 and *C*_*i*_ >0) or without initial confirmed cases (*C*_0*i*_>0 and *C*_*i*_ >0) in Stage One to identify the impact of the same mobility pattern on different kinds of cities. Statistical decisions were all made at a 5% level of significance using SPSS software.

### Estimating the impact of local travel restrictions on the epidemic spread

We assume, after the Wuhan lockdown, local travel restrictions in cities outside Hubei contributed to the epidemic control by influencing population mobility^10,16^. Data for three mobility patterns, except the population outflow from Wuhan, in two scenarios were obtained under assumptions. We assume the transmission conditions and virus characteristics in China would remain unchanged, and the best-fit models generated from the regression analysis were used to estimate the number of confirmed cases in cities outside Hubei based on the changed mobility data. The differences between the estimated cases and the confirmed cases are caused by the implementation of local travel restrictions, implying they have an impact on control of the epidemic.

To simulate the scenario of no travel restrictions in cites outside Hubei (Scenario 1), we used the 2019 Baidu data and assumed the mobility scale captured in 2019 would be similar to those of the equivalent time period during 2020^19^. All data before the Wuhan lockdown remained unchanged. Therefore, we adopted the mobility scale of 2019 and mobility direction of 2020 to simulate the population mobility after 23 January 2020, when there were no local travel restrictions. The daily population inflow from Hubei (excluding Wuhan), population inflow from cities outside Hubei, and intra-city population movement after 3 February 2019, aligned by the Chinese lunar calendar with 23 January 2020, were used as proxy mobility data for the no-local-travel-restrictions status in cities outside Hubei.

To understand the role of the relative timing imposition of local travel restrictions in other cities from the Wuhan lockdown, our Scenario 2 assumes more timely travel restrictions being imposed in cities outside Hubei. This scenario is only assumed to happen in Stage One, because local travel bans have fully implemented in Stage Two. The same models and methods were used as above, and the only change was the mobility data. Before 30 January, cities successively suspended their inter-provincial transport to and from Hubei (Supplementary Table 1), and restricted their inter-city and their intra-city population movement to varying degrees^6^. That is to say, the inter-city and intra-city population movements were all limited after 30 January. We assume all travel restrictions in cities outside Hubei were imposed at the same time as the Wuhan lockdown. Thus, the population outflow from Hubei excluding Wuhan solely considered those on 23 January. The population inflow from other cities and intra-city population movement from 31 January–6 February replaced those from 23–30 January as proxy mobility data for the timely travel restrictions status.

## Data Availability

We collated epidemiological data and mobility data from publicly available data sources. All the data sources we used are documented in the main text and supplementary tables.

## Contributors

LH and BX designed the experiments. LH and LZ collected data. LH analyzed data and wrote the first draft, and LY calculated the figures. LH, BX, SH, and PX interpreted the results and contributed the final manuscript. All authors approved the final version for submission.

